# General Anxiety Disorder-7 Questionnaire as a marker of low socioeconomic status and inequity

**DOI:** 10.1101/2021.12.21.21268205

**Authors:** Julio C. Nunes, Megan K. Carroll, Kenneth W. Mahaffey, Robert M. Califf, P. Murali Doraiswamy, Sarah Short, Svati H. Shah, Susan Swope, Donna Williams, Adrian F. Hernandez, David S. Hong

**Affiliations:** Stanford Center for Clinical Research, Stanford University School of Medicine, Stanford, CA, USA; Verily Life Sciences (Alphabet), San Francisco, CA, USA; Department of Psychiatry and Behavioral Sciences, Duke University School of Medicine, Durham, NC, USA; Duke Institute for Brain Sciences, Duke University School of Medicine, Durham, NC, USA; Duke Molecular Physiology Institute, Duke University School of Medicine, Durham, NC, USA; Division of Cardiology, Department of Medicine, Duke University School of Medicine, Durham, NC, USA; Duke Clinical Research Institute, Duke University School of Medicine, Durham, NC, USA; Department of Psychiatry and Behavioral Sciences, Stanford University School of Medicine, Stanford, CA, USA

**Keywords:** Generalized Anxiety Disorder-7, measures of health and disease, effective clinical intervention, social determinants of health

## Abstract

**Background:** The General Anxiety Disorder-7 (GAD-7) questionnaire is a standard tool used for screening and follow-up of patients with Generalized Anxiety Disorder (GAD). Although it is generally accepted that anxiety correlates with clinical and psychosocial stressors, precise quantitative data is limited on the relations among GAD-7, traditional biomarkers, and other measures of health. Even less is known about how GAD-7 relates to race, ethnicity, and socioeconomic status (SES).We determined how multiple demographic and socioeconomic data correlate with the participants’ GAD-7 results when compared with laboratory, physical function, clinical, and other biological markers.

**Methods:** The Project Baseline Health Study (BHS) is a prospective cohort of adults representing several populations in the USA. We analyzed a deeply phenotyped group of 2502 participants from that study. Measures of interest included: clinical markers or history of medical diagnoses; physical function markers including gait, grip strength, balance time, daily steps, and echocardiographic parameters; psychometric measurements; activities of daily living; socioeconomic characteristics; and laboratory results.

**Results:** Higher GAD-7 scores were associated with female sex, younger age, and Hispanic ethnicity. Measures of low SES were also associated with higher scores, including unemployment, income ≤$25,000, and ≤12 years of education. After adjustment for 166 demographic, clinical, laboratory, and symptom characteristics, unemployment and overall higher SES risk scores were highly correlated with anxiety scores. Protective factors included Black race and older age.

**Limitations:** Correlations identified in this cross-sectional study cannot be used to infer causal relationships; further, we were not able to account for possible use of anxiety treatments by study participants.

**Conclusions:** These findings highlight the importance of understanding anxiety as a biopsychosocial entity. Clinicians and provider organizations need to consider both the physical manifestations of the disorder and their patients’ social determinants of health when considering treatment pathways and designing interventions.

## INTRODUCTION

Anxiety disorders are among the most common psychiatric diagnoses, with a lifetime prevalence of 32% in the United States.^1^ Worldwide, the World Health Organization estimates that about 300 million people are affected.^2^ In the United States, anxiety disorders may contribute $110 million to $600 million per million inhabitants in indirect costs to the health care system.^3^ They also carry a large individual cost-burden due to their high prevalence, and may lead to school absences, work underperformance, socialization impairment, and overall disability.^4^ Anxiety is closely related to multiple chronic disorders, such as cardiovascular disease, diabetes mellitus type 2, hypertension, arthritis, and chronic obstructive pulmonary disease.^5-7^

Generalized Anxiety Disorder (GAD) is a chronic recurring subtype of anxiety, defined by persistent, excessive, difficult-to-control, and intrusive thoughts that may manifest comorbidly with somatic symptoms, sleep disturbances, and mental exhaustion. It frequently co-occurs with other psychiatric conditions, with at least 60% of the patients reporting having another disorder, mainly major depression, substance use disorder, attention deficit-hyperactivity disorder, and other types of anxiety.^1^

Spitzer et al. created the first standardized and widely available measurement tool for symptoms of GAD, the General Anxiety Disorder-7 (GAD7).^8^ Consisting of 7 questions scored from 0 to 3, this tool is easily usable by clinicians of all specialties and levels. It has since been validated in primary care, general population, outpatient, and inpatient psychiatric settings.^9-11^ The tool’s 7 components assess for 1) feeling nervous or on edge, 2) capacity to control worries and thoughts, 3) difficulty relaxing, 4) excessive worry about multiple themes, 5) restlessness, 6) irritability, and 7) constant worry or fear that unpleasant things will soon happen.

Several studies have examined possible correlations between higher degrees of anxiety and changes in laboratory measures, particularly biomarkers and peptides, related to serotonergic (e.g. 5-HT reuptake binding density, 5-HT plasma concentration), noradrenergic (e.g., platelet alpha-2 adrenergic peripheral binding density), and GABAergic (e.g., lymphocytes peripheral BDZ binding sites) systems.^12-17^ Oxidative and immunological systems are also of interest, and previous studies have shown increased levels of inflammatory markers and reactive oxygen radicals in patients experiencing intense degrees of anxiety.^18-22^ Vismara et al. recently reviewed and summarized the relationship between anxiety disorders and these biomarkers.^23^ However, despite these significant advances in understanding the physiology of anxiety, little has been published about how it correlates with wide-ranging population health markers, demographic characteristics, and social determinants of health.

The Baseline Health Study (BHS)^24^ is an ongoing prospective cohort study that aims to determine overall health biomarkers, biopsychosocial status, and demographics of a group of participants whose sex, race, and ethnicities are representative of the broader US adult population. In this study patients are deeply phenotyped, meaning that various demographic, laboratory, clinical, physical function, psychometric, and imaging findings are obtained annually during longitudinal follow-up visits. The BHS represents a unique opportunity to assess multiple types of anxiety correlates.

In this study, we focus on how multiple demographic and socioeconomic data correlate with GAD-7 results when compared with laboratory, physical function, clinical, and other biological markers. Our goal is to understand what role socioeconomic status (SES) and multiple other determinants of health may play in the biopsychosocial approach of anxiety disorders. Although some of the individual correlations have previously been reported, we add to the current knowledge by demonstrating the full range of these correlations in a single population, due to the comprehensive nature of data collection in the BHS.

## METHODS

### The Baseline Health Study

The Baseline Health Study is an ongoing multicenter longitudinal observational study. Multimodal data, including 166 different demographic, clinical, imaging, and laboratory measurements, were collected for 2502 adult participants. The BHS design has been previously described, including its data collection plan, inclusion and exclusion criteria, institutional review board regulation, and other main components.^24^

Study participants were recruited and enrolled via an online registration platform. The deeply phenotyped cohort used for this analysis was selected through an algorithm designed to produce a cohort that corresponds proportionately to U.S. adult population demographic data. Persons with various health statuses were included in the study, ranging from healthy controls to patients living with advanced disease. Due to the BHS methodology, the sampling was designed to over-represent persons with heart disease or cancer.

For this study, we performed a cross-sectional analysis that encompasses information of the baseline visit, when GAD-7 and the majority of the clinical data were collected. The GAD-7 was completed either at the in-person visit or digitally after the visit.

The BHS is funded by Verily Life Sciences (San Francisco, CA) and conducted by a collaborative group including Stanford University (Stanford, CA), Duke University (Durham, NC), and the California Health and Longevity Institute (Westlake Village, CA). The enrolling sites locations are Palo Alto, CA; Durham, NC; Kannapolis, NC; and Los Angeles, CA.

### Statistical Analysis

Demographic and clinical characteristics were described across the five GAD-7 severity categories (0, 1-4, 5-9, 10-14, 15+), separately for women and men, in the population of participants who completed the GAD-7 survey at baseline. To test associations, the Cochrane-Armitage trend test was used for binary variables, while the Spearman rank correlation test was used for continuous variables. Given the exploratory nature of the study, we did not adjust for multiple tests. Data were considered non-missing if they were collected within 200 days of a participants’ baseline in-person study visit.

### SES Risk Score

Socioeconomic status variables were combined into a single “SES risk score” variable that was used in subsequent LASSO regressions and on Figure 2. The variable was a sum score of 5 individual items coded as 1/0: 1) reported high school education or less, 2) reported income < $25,000, 3) marital status reported as unmarried, 4) employment status reported as not working, and 5) insurance status reported as uninsured. A higher score was considered as having “higher risk” SES.

### Imputation of Missing Data

All categorical predictors were converted into indicator variables via dummy coding. Multiple imputation by chained equations (MICE) was used to address missing data, as LASSO regression techniques require complete data on the input dataset. MICE assumes that all data are missing at random (MAR), meaning that the probability of a data point being missing depends only on observed data. The process generally consists of 4 steps: 1) a simple imputation (i.e., mean) is done for each missing value and used as placeholder, 2) placeholder imputations are set back to missing for a single “target” variable *X*_*i*_, 3) *X*_*i*_ becomes the dependent variable and is regressed on all other variables in the imputation model, and 4) missing values in *X*_*i*_ are replaced with predicted values from the model in step 3, and are then used in following regression models on variables with missing data. Steps 3-4 employ predictive mean matching in order to calculate the missing values of both continuous and categorical target variables; this ensures that missing data are assigned a value that has already been observed in the data. Steps 2-4 are repeated for each variable with missing data until all data are complete, a process that constitutes 1 “cycle.” A preset number of cycles were completed, at which point a final imputed dataset was computed. This process was then repeated for a prespecified number of imputed datasets.^25^

### LASSO Regression

LASSO procedures addressing multiply imputed data by using group methods (MI-LASSO) were conducted to select models of baseline characteristics that may be predictive of GAD-7 score (logarithm of GAD-7 + 1). All variables were standardized to allow for meaningful comparison of coefficients. In the MI-LASSO method, an L_1_-norm (the sum of the absolute values of the regression coefficients) penalization is placed on the regression model; this penalizes the inclusion of additional variables, and forces the coefficients of variables that do not contribute to the model to zero. In the presence of multiply imputed data, LASSO can produce different results for each imputed dataset; MI-LASSO fits models on all datasets jointly while considering each set of estimated coefficients (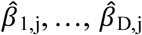, where D is the number of imputed datasets) as a group, and applying the group LASSO penalty to the model. Shrinkage of the coefficients is controlled by the tuning parameter (λ), which is optimized by selecting the model that minimizes the Bayesian Information Criterion (BIC).^26^

Analyses were performed using R v3.6.3 (R Foundation for Statistical Computing, Vienna, Austria). Data were imputed using the ‘MICE’ package v3.13.0.^27^ MI-LASSO regressions were run using the MI.LASSO function developed by Chen and Wang.^26^ Figures were created using the ‘ggplot2’ package v3.3.0.^28^

## RESULTS

A total of 2,453 participants completed baseline GAD-7 assessments, of whom 1,343 (55%) were female and 1,110 (45%) were male. Associations between GAD-7 scores and demographic data are shown in **Table 1**. Younger age, Hispanic ethnicity, and identifying as other race (defined as one of: Native Hawaiian or Pacific Islander, American Indian or Alaska Native, or Other race) were associated with higher GAD-7 scores for both men and women. The study’s population racial-ethnic distribution represents that of the US census, with the exception of Hispanic persons, who are underrepresented (11.3% of the study population compared with 18.5% according to 2019 estimates from the US Census).

**Table 1.**
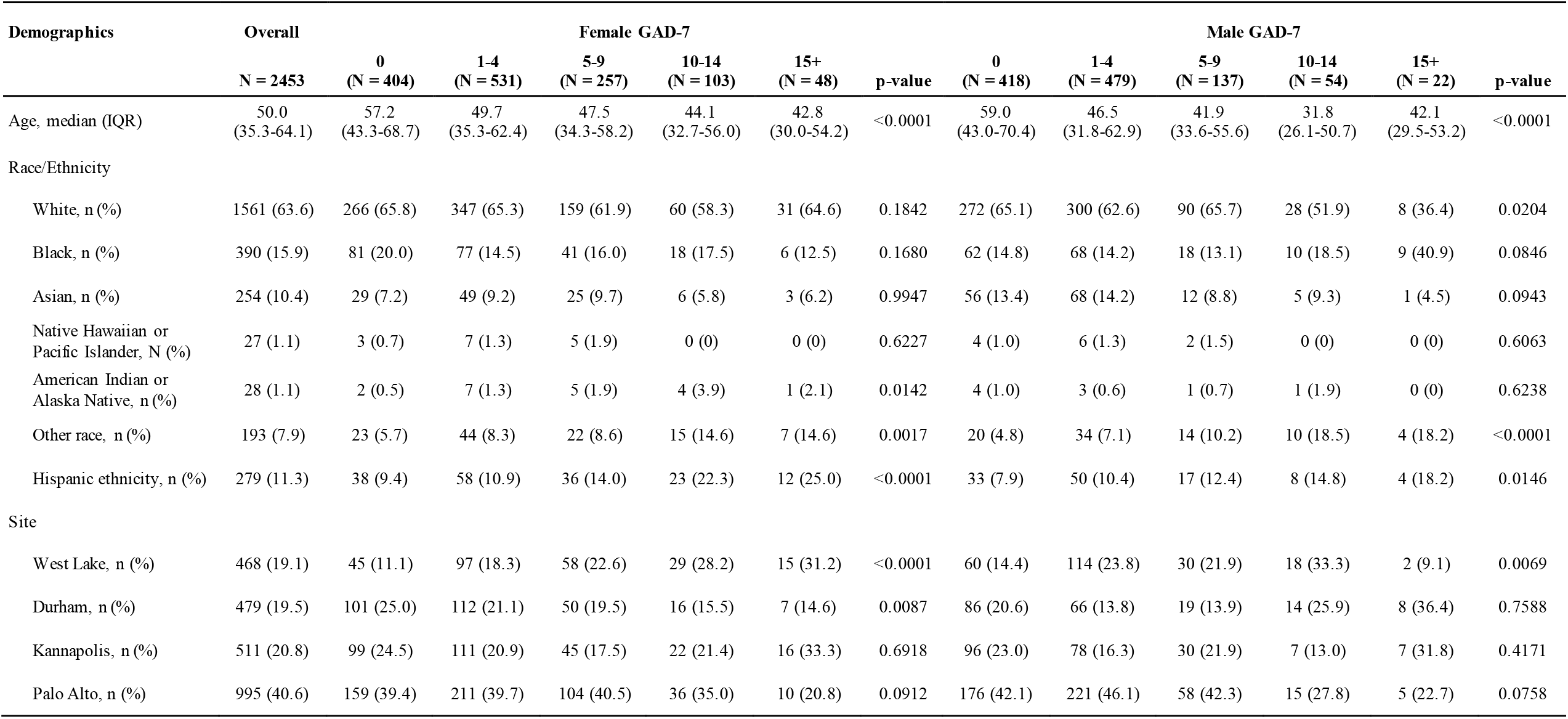
Summary of demographic characteristics across GAD-7 severity scores at baseline, stratified by sex. Data are presented as N (%) or median (IQR). P-values for trend were calculated with the use of Spearman Correlation or Cochrane-Armitage tests, where appropriate.

| Demographics | Overall<br>N = 2453 | Female GAD-7 |  |  |  |  |  | Male GAD-7 |  |  |  |  |  |
| --- | --- | --- | --- | --- | --- | --- | --- | --- | --- | --- | --- | --- | --- |
|  |  | 0<br>(N = 404) | 1-4<br>(N = 531) | 5-9<br>(N = 257) | 10-14<br>(N = 103) | 15+<br>(N = 48) | p-value | 0<br>(N = 418) | 1-4<br>(N = 479) | 5-9<br>(N = 137) | 10-14<br>(N = 54) | 15+<br>(N = 22) | p-value |
| Age, median (IQR) | 50.0<br>(35.3-64.1) | 57.2<br>(43.3-68.7) | 49.7<br>(35.3-62.4) | 47.5<br>(34.3-58.2) | 44.1<br>(32.7-56.0) | 42.8<br>(30.0-54.2) | <0.0001 | 59.0<br>(43.0-70.4) | 46.5<br>(31.8-62.9) | 41.9<br>(33.6-55.6) | 31.8<br>(26.1-50.7) | 42.1<br>(29.5-53.2) | <0.0001 |
| Race/Ethnicity |  |  |  |  |  |  |  |  |  |  |  |  |  |
| White, n (%) | 1561 (63.6) | 266 (65.8) | 347 (65.3) | 159 (61.9) | 60 (58.3) | 31 (64.6) | 0.1842 | 272 (65.1) | 300 (62.6) | 90 (65.7) | 28 (51.9) | 8 (36.4) | 0.0204 |
| Black, n (%) | 390 (15.9) | 81 (20.0) | 77 (14.5) | 41 (16.0) | 18 (17.5) | 6 (12.5) | 0.1680 | 62 (14.8) | 68 (14.2) | 18 (13.1) | 10 (18.5) | 9 (40.9) | 0.0846 |
| Asian, n (%) | 254 (10.4) | 29 (7.2) | 49 (9.2) | 25 (9.7) | 6 (5.8) | 3 (6.2) | 0.9947 | 56 (13.4) | 68 (14.2) | 12 (8.8) | 5 (9.3) | 1 (4.5) | 0.0943 |
| Native Hawaiian or Pacific Islander, N (%) | 27 (1.1) | 3 (0.7) | 7 (1.3) | 5 (1.9) | 0 (0) | 0 (0) | 0.6227 | 4 (1.0) | 6 (1.3) | 2 (1.5) | 0 (0) | 0 (0) | 0.6063 |
| American Indian or Alaska Native, n (%) | 28 (1.1) | 2 (0.5) | 7 (1.3) | 5 (1.9) | 4 (3.9) | 1 (2.1) | 0.0142 | 4 (1.0) | 3 (0.6) | 1 (0.7) | 1 (1.9) | 0 (0) | 0.6238 |
| Other race, n (%) | 193 (7.9) | 23 (5.7) | 44 (8.3) | 22 (8.6) | 15 (14.6) | 7 (14.6) | 0.0017 | 20 (4.8) | 34 (7.1) | 14 (10.2) | 10 (18.5) | 4 (18.2) | <0.0001 |
| Hispanic ethnicity, n (%) | 279 (11.3) | 38 (9.4) | 58 (10.9) | 36 (14.0) | 23 (22.3) | 12 (25.0) | <0.0001 | 33 (7.9) | 50 (10.4) | 17 (12.4) | 8 (14.8) | 4 (18.2) | 0.0146 |
| Site |  |  |  |  |  |  |  |  |  |  |  |  |  |
| West Lake, n (%) | 468 (19.1) | 45 (11.1) | 97 (18.3) | 58 (22.6) | 29 (28.2) | 15 (31.2) | <0.0001 | 60 (14.4) | 114 (23.8) | 30 (21.9) | 18 (33.3) | 2 (9.1) | 0.0069 |
| Durham, n (%) | 479 (19.5) | 101 (25.0) | 112 (21.1) | 50 (19.5) | 16 (15.5) | 7 (14.6) | 0.0087 | 86 (20.6) | 66 (13.8) | 19 (13.9) | 14 (25.9) | 8 (36.4) | 0.7588 |
| Kannapolis, n (%) | 511 (20.8) | 99 (24.5) | 111 (20.9) | 45 (17.5) | 22 (21.4) | 16 (33.3) | 0.6918 | 96 (23.0) | 78 (16.3) | 30 (21.9) | 7 (13.0) | 7 (31.8) | 0.4171 |
| Palo Alto, n (%) | 995 (40.6) | 159 (39.4) | 211 (39.7) | 104 (40.5) | 36 (35.0) | 10 (20.8) | 0.0912 | 176 (42.1) | 221 (46.1) | 58 (42.3) | 15 (27.8) | 5 (22.7) | 0.0758 |

Table 2 describes associations between markers of socioeconomic status and GAD-7 scores. Less education, lower income, unmarried status, not working for at least 1 year, and uninsured status were all associated with higher anxiety scores in both men and women. In general, higher GAD-7 scores were associated with lower SES across multiple types of measurement.

**Table 2.** Summary of socioeconomic status across GAD-7 severity scores at baseline, stratified by sex. Data are presented as N (%) or median (IQR). P-values for trend were calculated with the use of Spearman Correlation or Cochrane-Armitage tests, where appropriate.

| Socioeconomic Status | Overall | Female GAD-7 |  |  |  |  |  | Male GAD-7 |  |  |  |  |  |
| --- | --- | --- | --- | --- | --- | --- | --- | --- | --- | --- | --- | --- | --- |
|  | N = 2453 | 0<br>(N = 404) | 1-4<br>(N = 531) | 5-9<br>(N = 257) | 10-14<br>(N = 103) | 15+<br>(N = 48) | p-value | 0<br>(N = 418) | 1-4<br>(N = 479) | 5-9<br>(N = 137) | 10-14<br>(N = 54) | 15+<br>(N = 22) | p-value |
| Education |  |  |  |  |  |  |  |  |  |  |  |  |  |
| High school or less, n (%) | 176 (7.2) | 28 (6.9) | 32 (6.0) | 17 (6.6) | 15 (14.6) | 9 (18.8) | 0.0024 | 25 (6.0) | 20 (4.2) | 15 (10.9) | 11 (20.4) | 4 (18.2) | <0.0001 |
| Some college, n (%) | 455 (18.5) | 70 (17.3) | 103 (19.4) | 71 (27.6) | 25 (24.3) | 11 (22.9) | 0.0076 | 65 (15.6) | 70 (14.6) | 24 (17.5) | 10 (18.5) | 6 (27.3) | 0.2192 |
| College, n (%) | 634 (25.8) | 120 (29.7) | 178 (33.5) | 68 (26.5) | 24 (23.3) | 11 (22.9) | 0.0721 | 95 (22.7) | 105 (21.9) | 24 (17.5) | 8 (14.8) | 1 (4.5) | 0.0192 |
| Graduate degree or higher, n (%) | 670 (27.3) | 123 (30.4) | 140 (26.4) | 58 (22.6) | 13 (12.6) | 4 (8.3) | <0.0001 | 144 (34.4) | 144 (30.1) | 37 (27.0) | 6 (11.1) | 1 (4.5) | <0.0001 |
| Income |  |  |  |  |  |  |  |  |  |  |  |  |  |
| < \$25,000, n (%) | 193 (7.9) | 24 (5.9) | 40 (7.5) | 34 (13.2) | 20 (19.4) | 9 (18.8) | <0.0001 | 20 (4.8) | 24 (5.0) | 6 (4.4) | 10 (18.5) | 6 (27.3) | <0.0001 |
| \$25,000-50,000, n (%) | 256 (10.4) | 43 (10.6) | 52 (9.8) | 39 (15.2) | 15 (14.6) | 9 (18.8) | 0.0186 | 34 (8.1) | 44 (9.2) | 14 (10.2) | 6 (11.1) | 0 (0.0) | 0.7462 |
| \$50,000-100,000, n (%) | 493 (20.1) | 86 (21.3) | 142 (26.7) | 53 (20.6) | 26 (25.2) | 6 (12.5) | 0.5255 | 70 (16.7) | 77 (16.1) | 24 (17.5) | 7 (13.0) | 2 (9.1) | 0.4488 |
| \$100,000-150,000, n (%) | 308 (12.6) | 57 (14.1) | 61 (11.5) | 28 (10.9) | 2 (1.9) | 2 (4.2) | 0.0006 | 64 (15.3) | 71 (14.8) | 19 (13.9) | 2 (3.7) | 2 (9.1) | 0.0678 |
| \$150,000-200,000, n (%) | 197 (8.0) | 35 (8.7) | 50 (9.4) | 16 (6.2) | 5 (4.9) | 2 (4.2) | 0.0643 | 43 (10.3) | 30 (6.3) | 12 (8.8) | 3 (5.6) | 1 (4.5) | 0.1106 |
| > \$200,000, n (%) | 358 (14.6) | 64 (15.8) | 77 (14.5) | 27 (10.5) | 8 (7.8) | 3 (6.2) | 0.0028 | 79 (18.9) | 75 (15.7) | 20 (14.6) | 5 (9.3) | 0 (0.0) | 0.0077 |
| Marital Status |  |  |  |  |  |  |  |  |  |  |  |  |  |
| Married, n (%) | 1054 (43.0) | 179 (44.3) | 242 (45.6) | 97 (37.7) | 31 (30.1) | 11 (22.9) | 0.0002 | 232 (55.5) | 200 (41.8) | 44 (32.1) | 13 (24.1) | 5 (22.7) | <0.0001 |
| Divorced, n (%) | 180 (7.3) | 42 (10.4) | 49 (9.2) | 30 (11.7) | 12 (11.7) | 4 (8.3) | 0.7771 | 20 (4.8) | 15 (3.1) | 4 (2.9) | 2 (3.7) | 2 (9.1) | 0.7563 |
| Formerly in long term relationship, n (%) | 97 (4.0) | 12 (3.0) | 22 (4.1) | 11 (4.3) | 3 (2.9) | 0 (0.0) | 0.9021 | 16 (3.8) | 24 (5.0) | 5 (3.6) | 3 (5.6) | 1 (4.5) | 0.6686 |
| Living together, n (%) | 205 (8.4) | 26 (6.4) | 46 (8.7) | 31 (12.1) | 9 (8.7) | 8 (16.7) | 0.0084 | 23 (5.5) | 34 (7.1) | 19 (13.9) | 8 (14.8) | 1 (4.5) | 0.0049 |
| Never in long term relationship, n (%) | 255 (10.4) | 38 (9.4) | 60 (11.3) | 22 (8.6) | 13 (12.6) | 9 (18.8) | 0.1884 | 26 (6.2) | 54 (11.3) | 21 (15.3) | 9 (16.7) | 3 (13.6) | 0.0004 |

**Table 2. Summary of socioeconomic status across GAD-7 severity scores at baseline, stratified by sex.** Data are presented as N (%) or median (IQR). P-values for trend were calculated with the use of Spearman Correlation or Cochran-Armitage tests, where appropriate.
| Socioeconomic Status | Overall | Female GAD-7 |  |  |  |  |  | Male GAD-7 |  |  |  |  |  |
| --- | --- | --- | --- | --- | --- | --- | --- | --- | --- | --- | --- | --- | --- |
|  | N = 2453 | 0<br>(N = 404) | 1-4<br>(N = 531) | 5-9<br>(N = 257) | 10-14<br>(N = 103) | 15+<br>(N = 48) | p-value | 0<br>(N = 418) | 1-4<br>(N = 479) | 5-9<br>(N = 137) | 10-14<br>(N = 54) | 15+<br>(N = 22) | p-value |
| Separated, n (%) | 48 (2.0) | 9 (2.2) | 6 (1.1) | 14 (5.4) | 5 (4.9) | 1 (2.1) | 0.0378 | 6 (1.4) | 4 (0.8) | 3 (2.2) | 0 (0.0) | 0 (0.0) | 0.8138 |
| Widowed, n (%) | 75 (3.1) | 29 (7.2) | 23 (4.3) | 5 (1.9) | 3 (2.9) | 1 (2.1) | 0.0025 | 3 (0.7) | 9 (1.9) | 2 (1.5) | 0 (0.0) | 0 (0.0) | 0.4172 |
| Employment Status |  |  |  |  |  |  |  |  |  |  |  |  |  |
| Employed for wages, n (%) | 1147 (46.8) | 189 (46.8) | 273 (51.4) | 121 (47.1) | 40 (38.8) | 20 (41.7) | 0.1892 | 175 (41.9) | 228 (47.6) | 71 (51.8) | 23 (42.6) | 7 (31.8) | 0.4526 |
| Homemaker, n (%) | 67 (2.7) | 13 (3.2) | 25 (4.7) | 18 (7.0) | 5 (4.9) | 4 (8.3) | 0.0372 | 0 (0.0) | 0 (0.0) | 1 (0.7) | 1 (1.9) | 0 (0.0) | 0.0128 |
| Unable to work, n (%) | 65 (2.6) | 1 (0.2) | 11 (2.1) | 11 (4.3) | 13 (12.6) | 4 (8.3) | <0.0001 | 5 (1.2) | 8 (1.7) | 5 (3.6) | 4 (7.4) | 3 (13.6) | <0.0001 |
| Not working ≥ 1 year, n (%) | 49 (2.0) | 5 (1.2) | 11 (2.1) | 8 (3.1) | 4 (3.9) | 2 (4.2) | 0.0281 | 4 (1.0) | 7 (1.5) | 6 (4.4) | 1 (1.9) | 1 (4.5) | 0.0279 |
| Retired, n (%) | 457 (18.6) | 106 (26.2) | 104 (19.6) | 25 (9.7) | 7 (6.8) | 5 (10.4) | <0.0001 | 121 (28.9) | 72 (15.0) | 12 (8.8) | 5 (9.3) | 0 (0.0) | <0.0001 |
| Student, n (%) | 56 (2.3) | 9 (2.2) | 11 (2.1) | 11 (4.3) | 2 (1.9) | 1 (2.1) | 0.5223 | 5 (1.2) | 13 (2.7) | 2 (1.5) | 2 (3.7) | 0 (0.0) | 0.2892 |
| Not working < 1 year, n (%) | 58 (2.4) | 11 (2.7) | 9 (1.7) | 7 (2.7) | 5 (4.9) | 1 (2.1) | 0.4999 | 9 (2.2) | 11 (2.3) | 4 (2.9) | 1 (1.9) | 0 (0.0) | 0.8428 |
| Self-employed, n (%) | 254 (10.4) | 34 (8.4) | 41 (7.7) | 35 (13.6) | 14 (13.6) | 2 (4.2) | 0.1543 | 46 (11.0) | 61 (12.7) | 14 (10.2) | 4 (7.4) | 3 (13.6) | 0.7879 |
| Health Insurance Status |  |  |  |  |  |  |  |  |  |  |  |  |  |
| Insured, n (%) | 1809 (73.7) | 321 (79.5) | 436 (82.1) | 192 (74.7) | 67 (65.0) | 32 (66.7) | 0.0003 | 315 (75.4) | 313 (65.3) | 93 (67.9) | 33 (61.1) | 7 (31.8) | <0.0001 |
| Uninsured, n (%) | 110 (4.5) | 17 (4.2) | 15 (2.8) | 20 (7.8) | 7 (6.8) | 3 (6.2) | 0.0391 | 11 (2.6) | 24 (5.0) | 7 (5.1) | 2 (3.7) | 4 (18.2) | 0.0083 |
| Smoking Status |  |  |  |  |  |  |  |  |  |  |  |  |  |
| Current smoker, n (%) | 331 (13.5) | 34 (8.4) | 56 (10.5) | 46 (17.9) | 25 (24.3) | 14 (29.2) | <0.0001 | 43 (10.3) | 62 (12.9) | 23 (16.8) | 17 (31.5) | 11 (50.0) | <0.0001 |
| Former smoker, n (%) | 539 (22.0) | 73 (18.1) | 119 (22.4) | 55 (21.4) | 21 (20.4) | 8 (16.7) | 0.6857 | 109 (26.1) | 105 (21.9) | 31 (22.6) | 11 (20.4) | 7 (31.8) | 0.4639 |
| Nonsmoker, n (%) | 1583 (64.5) | 297 (73.5) | 356 (67.0) | 156 (60.7) | 57 (55.3) | 26 (54.2) | <0.0001 | 266 (63.6) | 312 (65.1) | 83 (60.6) | 26 (48.1) | 4 (18.2) | 0.0005 |

Five datasets were imputed from the observed data using 5 cycles per imputation; the 5 datasets were simultaneously entered in the MI-LASSO model. Results from the full MI-LASSO regression models, which included 339 total predictors and was stratified by sex, are presented in Figure 1. Predictors classically described as somatic symptoms of anxiety per DSM-V criteria (e.g., headaches) were not included in the final model to allow for the observation of factors (e.g., symptoms of other medical conditions; sociodemographic data) not yet established as components of the syndrome of anxiety. A full list of variables entered into the MI-LASSO regression is available in Appendix A. In both men and women, memory change was the strongest positive predictor of GAD-7 severity. In women, the coefficient was more than twice that of the next strongest positive predictor (shortness of breath); in men, the coefficient was more than 3 times that of the next strongest positive predictor (muscle or joint pain). For women, we also observed positive associations between GAD-7 severity and other physical health-related symptoms (e.g., coughing up sputum, lightheadedness, diarrhea, and constipation), absolute monocyte levels, reporting Other race, and SES score. There were negative relationships between GAD-7 severity and both age and Black race. In men, there were positive associations between GAD-7 and other physical function measures including shortness of breath with exercise, joint pain or swelling, shortness of breath, and constipation. Similar to findings observed in women, GAD-7 severity decreased as age increased in men.

**Figure 1.**
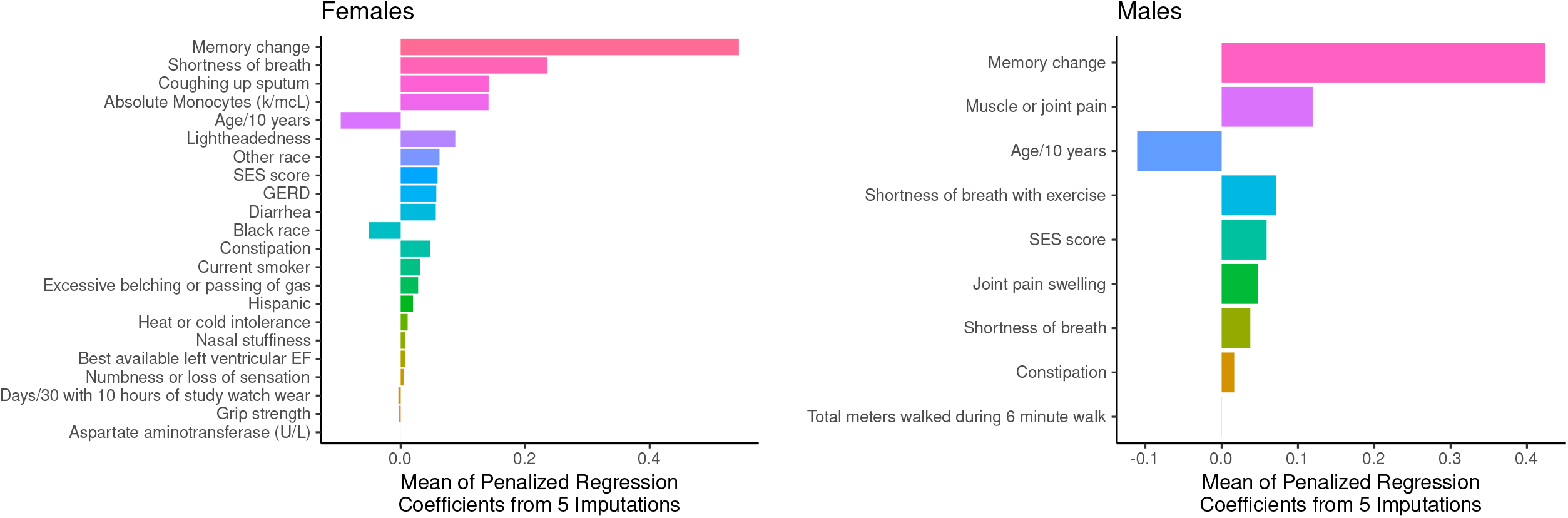
Factors associated with GAD-7 score after MI-LASSO regression. The mean of the MI-LASSO regression coefficients from 5 imputed datasets that predict GAD-7 severity are presented for both male and female study participants. Coefficients are sorted in descending order of magnitude.

Figure 2 presents a heat map figure of associations between SES risk scores and GAD-7 severity, stratified by race. Overall associations between SES risk score and GAD-7 severity were significant for all races. Black participants tended to have higher concentrations in the high-SES risk, high-GAD-7 severity corner of the table; as SES risk score increased, the likelihood of having a higher GAD-7 severity score also increased. Black participants were also most likely to have an SES score of at least 3. White participants had higher concentrations in the low-SES risk, low GAD-7 corner of the table; although there was a similar trend toward higher GAD-7 scores as SES risk score increased, white participants had lower SES risk scores overall. Asian participants were more likely to have low SES risk, and few participants (n=13) had a GAD-7 severity of at least 10. There was no clear trend between SES risk score and GAD-7 severity for those who reported other races, but those participants tended to have lower SES risk scores overall.

**Figure 2.**
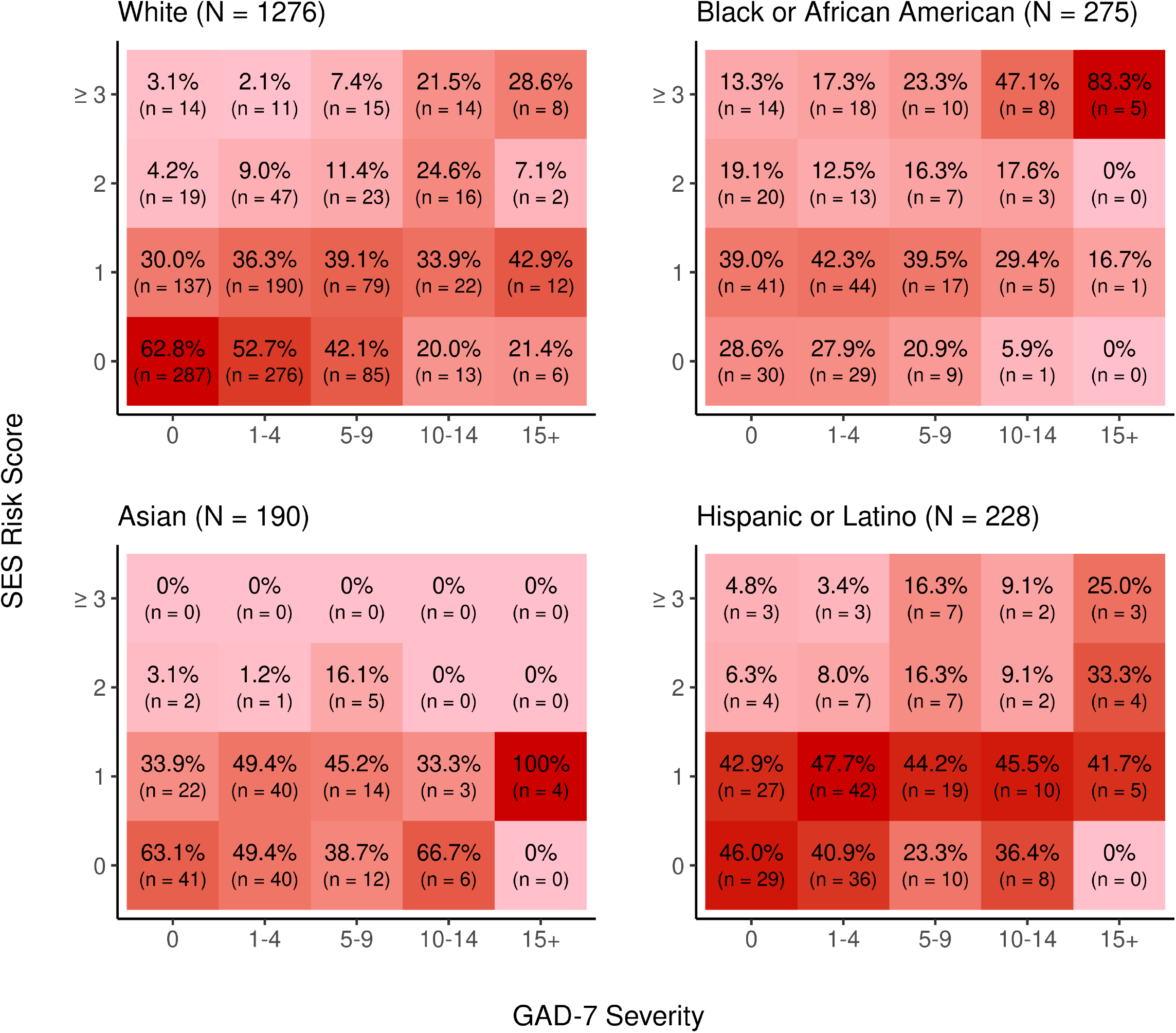
Associations between socioeconomic status (SES) and GAD-7 score, stratified by race. Each panel describes the interaction of GAD-7 severity and SES risk score within each of 4 racial groupings. Column percentages were calculated as the counts within an individual cell divided by the total count of participants given that cells’ GAD-7 severity score. Individual cells were shaded based on the column percentage; darker cells indicate a greater column percentage. Fisher Exact Tests for count data with simulated p-value were performed within each race; significant overall associations between SES risk score and GAD-7 severity for all races (White: p < 0.001; Black or African American: p = 0.012; Asian: p = 0.010; Other: p = 0.02

## DISCUSSION

Our study enrolled a large, deeply phenotyped cohort that allowed us to assess a broad array of anxiety correlates. Our findings confirm and highlight previous literature data regarding GAD-7 and different measurements of SES. We emphasize the importance of using GAD-7 with psychosocial contextual knowledge, as it raises concerns not only about psychological symptoms, but also about broader social determinants of health. Although some of the individual correlations are expected, their assembly highlights the importance of the biopsychosocial model when managing mental health disorders.

Previous studies have demonstrated how low income, unemployment, fewer years of education, lack of insurance, and smoking correlate with higher levels of anxiety. Hinz et al. validated the psychometric properties of the GAD-7 in Germany and found that those social determinants of health were correlated with higher anxiety scores.^29^ Finegan et al. (2020) investigated the influence of socioeconomic deprivation and neighborhood violence on anxiety and found similar results but also observed that low neighborhood average income and high crime rates were likewise correlated with higher GAD-7 scores.^30^ Chen et al. reported similar findings in an analysis of 13,775 medical records.^31^

When designing interventions, clinicians and policy makers must consider socioeconomic issues, access to opportunities, and social context to better reduce anxiety. Different treatment modalities may be more effective if employed in a holistic approach that is sensitive to more than just biochemical matters or cognitive function. Furthermore, SES may limit the types of treatment and therapy patients may receive and thus, clinicians and health systems should be focused on innovative methods to enable people with anxiety to access the standard of care.^32,33^

Identifying as a person of Hispanic/Latinx ethnicity was correlated with higher GAD-7 scores, a result consistent with previously published findings. Factors that may contribute to this correlation include higher rates of unemployment, job insecurity, lack of access to health care, distance from family, use of English as a second language, and systemic racism and prejudice.^34,35^ Additionally, concerns about migration and visa status could lead certain ethnic groups to not enroll, which is reflected on the current underrepresentation of Latinx populations in our sample. We did not ask participants about their visa status and that is a limitation of this study. This aspect is important, as insecurity around residency status may also contribute to higher degrees of psychosocial distress among Hispanic populations.^36,37^

After adjustment for multiple factors, Black race was correlated with lower overall GAD-7 scores, a finding consistent with previous studies suggesting that Black individuals may have higher levels of resiliency when facing adversity.^38^ However, when focusing on the relationships between race, SES risk score, and GAD-7 score (Figure 2), Black persons with high SES risk mostly have higher scores, suggesting that poor SES may be a significant associated factor in the development of anxiety for that group. This contrasts with white participants, who had higher levels of anxiety distributed across all levels of SES. Nevertheless, this unexpected finding should be considered preliminary and warrants further investigation.

We also observed higher levels of anxiety among young unmarried women. These findings were previously reported in the literature and most recently updated during the COVID-19 pandemic. In studies of temporal trends and demographic disparities of mental health disorders, sex, age, and marital status were predictors of higher deterioration of mental health status, particularly when stricter social isolation measures were in place.^39,40^

The BHS is longitudinally assessing a large cohort of patients for markers of social, biological, clinical, and behavioral function; once it is completed, we will have an opportunity to better understand these relationships across time. Additionally, by the end of the study, race- and ethnicity-specific analyses may allow a more nuanced understanding of how social determinants of health shape the profile of anxiety in specific populations. The representation of each demographic group is a priority for the BHS study and its final population pool will reflect every race/ethnicity per the US census. The BHS is also an opportunity to understand the validity of behavioral data collection in the context of digital health.

### Limitations

A number of limitations to this study should be noted. Cross-sectional studies cannot determine causality. Therefore, this study is capable of demonstrating anxiety correlations but does not attempt to explain basic underlying causal effects. In addition, we lack details about anxiety treatments, which could be an unaccounted modifying factor. In future longitudinal follow-up studies, we plan to thoroughly consider behavioral treatments and medications and their associations.

## CONCLUSION

In conclusion, higher GAD-7 scores were correlated with multiple measures across the spectrum of biological, clinical, behavioral and social measures. The relationship with SES is particularly striking, highlighting the importance of considering social determinants of health when designing interventions for anxiety. Focusing solely on the biological treatment of anxiety or the personal cognitive issues may have a limited effectiveness when compared with holistic approaches to care. Clinicians and policy makers are likely to be more effective if they are aware of social and contextual circumstances unique to the patient combined with social factors affecting groups. Additionally, when systems are set in place aiming to improve SES of individuals and populations, anxiety should be considered as a factor that may limit the capacity of these individuals to participate and respond to those interventions.

## Data Availability

The Baseline Study data will be available to qualified researchers for exploratory analysis after the data are adequately curated and initial planned primary manuscripts are written. Qualified external researchers will be able to apply through applications reviewed by the Proposal Review and Publications Committee and Scientific Executive Committee.

## Contributors

JCN drafted and wrote the paper and performed the analysis. MKC contributed data or analysis tools, performed the analysis, wrote the paper. KWM conceived and designed the analysis, reviewed the paper. RMC conceived and designed the analysis, reviewed the paper. PMD conceived and designed the analysis, reviewed the paper. S Short contributed data or analysis tools, performed the analysis, reviewed the paper. SHS contributed data or analysis tools, reviewed the paper. S Swope collected the data, reviewed the paper. DW collected the data, reviewed the paper. AFH conceived and designed the analysis, reviewed the paper. DSH conceived and designed the analysis, wrote and reviewed the paper, coordinated writing and analysis efforts.

## Declaration of Competing Interest

None.

## Disclosures

Dr. Califf is an employee of Verily Life Sciences and Google Health; he also serves as an advisory board/science advisory board member for Human Health and Potential (Singapore), Basking Biosciences, Launch and Scale Speedometer, and Medicxi Ventures. He is also a board member for AMYRIAD Pharma, Centessa Pharmaceuticals, Clinetic, One Fifteen, Portola, and Cytokinetics. Ms. Carroll and Ms. Short are employees of Verily Life Sciences. Dr. Hernandez notes external relationships with Amgen, AstraZeneca, Bayer, Biofourmis Singapore, Boehringer Ingelheim, Boston Scientific, Cytokinetics, Eidos Therapeutics, Eli Lilly, Intercept, Luitpold Pharmaceuticals, Merck, MyoKardia, Novartis, Pfizer, Relypsa, and Verily. Dr. Doraiswamy notes external relationships with Advera Health, Apollo Hospitals, Danone, Evidation Health, Live Love Laugh, Lumos Labs, MarvelBiome, Neuroglee, Transposon Therapeutics, Turtle Shell Technologies, UMethod, VitaKey, and Vivly. Dr. Shah reports external relationships with American Heart Association, AstraZeneca, Baylis Medical, Biosense Webster, Cardivia Medical, Lilly, McGraw-Hill Publishing, the National Institutes of Health, NewPace, and Project Baseline, LLC. Dr. Mahaffey’s financial disclosures can be viewed at http://med.stanford.edu/profiles/kenneth-mahaffey.

## Acknowledgments

Baseline Health Study Team: American Society of Clinical Oncology, Alexandria, VA, USA: Richard L. Schilsky. Duke University, School of Medicine, Durham, NC, USA: Jennifer Allen, MaryAnn Anderson, Kevin Anstrom, Lucus Araujo, Kristine Arges, Kaveh Ardalan, Bridget Baldwin, Suresh Balu, Mustafa R. Bashir, Manju Bhapkar, Robert Bigelow, Tanya Black, Rosalia Blanco, Gerald Bloomfield, Durga Borkar, Leah Bouk, Ebony Boulware, Nikki Brugnoni, Erin Campbell, Paul Campbell, Larry Carin, Tammy Jo Cassella, Tina Cates, Ranee Chatterjee Montgomery, Victoria Christian, John Choong, Michael Cohen-Wolkowiez, Elizabeth Cook, Scott Cousins, Ashley Crawford, Nisha Datta, Melissa Daubert, James Davis, Jillian Dirkes, Isabelle Doan, Marie Dockery, P. Murali Doraiswamy, Pamela S. Douglas, Shelly Duckworth, Ashley Dunham, Gary Dunn, Ryan Ebersohl, Julie Eckstrand, Vivienne Fang, April Flora, Emily Ford, Lucia Foster, Elizabeth Fraulo, John French, Geoffrey S. Ginsburg, Cindy Green, Latoya Greene, Jeffrey Guptill, Donna Hamel, Jennifer Hamill, Chris Harrington, Rob Harrison, Lauren Hedges, Brooke Heidenfelder, Adrian F. Hernandez, Cindy Heydary, Tim Hicks, Lina Hight, Deborah Hopkins, Erich S. Huang, Grace Huh, Jillian Hurst, Kelly Inman, Gemini Janas, Glenn Jaffee, Janace Johnson, Tiffanie Keaton, Michel Khouri, Daniel King, Jennifer Korzekwinski, Lynne H. Koweek, Anthony Kuo, Lydia Kwee, Dawn Landis, Rachele Lipsky, Desiree Lopez, Carolyn Lowry, Kelly Marcom, Keith Marsolo, Paige McAdams, Shannon McCall,, Robert McGarrah, John McGugan, Dani Mee, Sabrena Mervin-Blake, Prithu Mettu, Mathias Meyer, Justin Meyers, Calire N. Miller, Rebecca Moen, Lawrence H. Muhlbaier, Michael Murphy, Ben Neely, L. Kristin Newby, Jayne Nicoldson, Hoang Nguyen, Maggie Nguyen, Lori O’Brien, Sumru Onal, Jeremey O’Quinn, David Page, Neha J. Pagidipati, Kishan Parikh, Sarah R. Palmer, Bray Patrick-Lake, Brenda Pattison, Michael Pencina, Eric D. Peterson, Jon Piccini, Terry Poole, Tom Povsic, Alicia Provencher, Dawn Rabineau, Annette Rich, Susan Rimmer, Fides Schwartz, Angela Serafin, Nishant Shah, Svati Shah, Kelly Shields, Steven Shipes, Peter Shrader, Jon Stiber, Lynn Sutton, Geeta Swamy, Betsy Thomas, Sandra Torres, Debara Tucci, Anthony Twisdale, Susan A. Whitney, Robin Williamson, Lauren Wilverding, Charlene A. Wong, Lisa Wruck. Ellen Young Gemini Group, USA: Jane Perlmutter. Health Collaboratory and Cancer 101, New York, NY, USA: Sarah Krug. Rare Dots, Inc., USA: S. Whitney Bowman-Zatzkin. Society of Participatory Medicine, USA: Sarah Krug. Stanford University, School of Medicine, Stanford, CA, USA: Themistocles Assimes, Vikram Bajaj, Maxwell Cheong, Millie Das, Manisha Desai, Alice C. Fan, Dominik Fleischmann, Sanjiv S. Gambhir, Garry Gold, Francois Haddad, David Hong, Curtis Langlotz, Yaping J. Liao, Rong Lu, Kenneth W. Mahaffey, David Maron, Rebecca McCue, Rajan Munshi, Fatima Rodriguez, Sumana Shashidhar, George Sledge, Susie Spielman, Ryan Spitler, Sue Swope, Donna Williams, Julio C. Nunes. University of Florida, College of Medicine, Gainesville, FL, USA: Carl J Pepine. University of Missouri, Children’s Mercy Hospital, Kansas City, MO, USA: John D Lantos. University of Texas, Dell Medical School, Austin, TX, USA: Michael Pignone. University of Washington, Department of Biostatistics, Seattle, WA, USA: Patrick Heagerty. Vanderbilt University, School of Medicine, Nashville, TN, USA: Laura Beskow, Gordon Bernard. Verily Inc., South San Francisco, CA, USA: Kelley Abad, Giulia Angi, Robert M. Califf, Lawrence Deang, Joy Huynh, Manway Liu, Cherry Mao, Michael Magdaleno, William J. Marks, Jr., Jessica Mega, David Miller, Nicole Ong, Darshita Patel, Vanessa Ridaura, Scarlet Shore, Sarah Short, Michelle Tran, Veronica Vu, Celeste Wong. Harvard University, School of Medicine, Boston, MA, USA: Robert C. Green. Google Inc., Mountain View, CA, USA: John Hernandez. California Health and Longevity Institute, Westlake Village, CA, USA: Jolene Benge, Gislia Negrete, Gelsey Sierra, Terry Schaack

## Funding

The Baseline Health Study and this analysis were funded by Verily Life Sciences, South San Francisco, CA. This analysis was funded by Verily Life Sciences (South San Francisco, CA) in partnership with Stanford University and Duke University.

## Role of the funding source

The employees from funding source contributed to the data analyses, interpretation, editing of the manuscript and are coauthors. The final decision to submit the manuscript was made by the academic authors.

## Notes

### Competing Interest Statement

The authors have declared no competing interest.

### Author Declarations

The Project Baseline Study (NCT03154346) was approved by a central IRB (Western IRB/WCG) and by the IRBs of all participating sites.

