## Appendix for "General Anxiety Disorder-7 Questionnaire as a marker of low socioeconomic status and inequity"

**Appendix A. List of candidate covariates for multivariable LASSO regression**

**A.1. Demographic and Baseline Characteristics**

| **Variable Name** | **Definition** | **Description of Derivation** |
| --- | --- | --- |
| age_at_enrollmentd10 | Age in years at baseline | (enrollment_date - DOB)/10 |
| female | Self-reported sex at birth | N/A |
| race_asian  race_black  race_other | Self-reported race | N/A |
| hispanic | Self-reported Hispanic ancestry (yes/no) | N/A |
| current_smoker  former_smoker | Self-reported smoking status at baseline | N/A |
| pack_years_smoked | Self-reported cigarettes smoked per day multiplied by years of regular smoking divided by 20 | cigarettes_per_day * years_regular_smoking / 20 |

**A.2. On Site Assessments - Vitals and Physical Health Metrics**

| **Variable Name** | **Definition** | **Description of Derivation** |
| --- | --- | --- |
| systolic_blood_pressure | Average of 2 systolic blood pressure readings (mmHg) measured at baseline | (sbp1_mmhg + sbp2_mmhg) / 2 |
| diastolic_blood_pressure | Average of 2 diastolic blood pressure readings (mmHg) measured at baseline | (dbp1_mmhg + dbp2_mmhg) / 2 |
| vs_pulse_bpm | Heart rate (bpm) measured at baseline | N/A |
| body_mass_index | Body weight (kg) divided by the square of height (cm) at baseline | weight_kg / (height_cm / 100)^2^ |
| vs_wc_cm | Waist circumference (cm) at baseline | N/A |
| vs_osat_pct | Oxygen saturation (%) at baseline | N/A |
| vs_rrate_bpm | Respiratory rate (bpm) at baseline | N/A |
| ankle_brachial_index | Minimum of left and right ankle brachial index at baseline, using the maximum of left dorsalis pedis pressure and left posterior tibial pressure, maximum of right dorsalis pedis pressure and right posterior tibial pressure, and maximum of right and left brachial systolic pressure | Minimum between:  Max (left_dp + left_pt) / max (right brachial_pressure + left_brachial_pressure) and max (right_dp + right_pt) / max (right brachial_pressure + left_brachial_pressure) |
| ratio_of_forced_expiratory_volume | Ratio of forced expiratory volume (first third of forced breath) and forced vital capacity at baseline | FEV1 / FVC |
| pperf_6mdis_m | Total meters walked during 6 minute walk distance test at baseline | N/A |
| ten_meter_walk_speed | Walking speed in meters per second using average of 3 fast walk trials during 10 meter walk test at baseline | 6 / mean (fw1_sec + fw2_sec + fw3_sec) |
| handgrip_dominant | Average of 3 hand grip trials of dominant hand in kilograms at baseline | mean (right1_kg + right2_kg + right3_kg) or mean (left1_kg + left2_kg + left3_kg) |
| single_legged_balance | Average of left and right leg trials in seconds during single-legged balance test at baseline | (slb_left_sec + slb_right_sec) / 2 |
| sitting_rising_score | Sum of sitting and rising scores during sitting-rising test at baseline | sitting_score + rising_score |
| pperf_30sscr | Number of stands during 30 second chair stand test at baseline | N/A |
| best_available_left_ventricular | Best available left ventricular EF from biplane, single and visual read data from resting echocardiogram at baseline | Biplane EF if available; if not then single plane EF; else visual EF |
| lvmassi | Left ventricular mass index from resting echocardiogram at baseline | N/A |
| ccs | Coronary calcium score from coronary calcium scan at baseline | N/A |

**A.3. On Site Assessments - Medical Conditions (60 most commonly reported), Symptoms (50 most commonly reported), and Allergies**

| **Variable Name** | **Definition** | **Description of Derivation** |
| --- | --- | --- |
| **Physical Health-Related Medical Conditions** | | |
| cc_oa | Self-reported osteoarthritis at baseline | N/A |
| cc_gerd | Self-reported gastroesophageal reflux disease at baseline | N/A |
| cc_htn | Self-reported hypertension at baseline | N/A |
| cc_asthma | Self-reported asthma at baseline | N/A |
| cc_cataracts | Self-reported cataracts at baseline | N/A |
| cc_hypercholesterolemia | Self-reported hypercholesterolemia at baseline | N/A |
| cc_dm2 | Self-reported type II diabetes at baseline |  |
| cc_sleepapnea | Self-reported sleep apnea at baseline | N/A |
| cc_colonpolyps | Self-reported colon polyps at baseline | N/A |
| cc_pneumonia | Self-reported pneumonia | N/A |
| cc_hypothyroidism | Self-reported hypothyroidism at baseline | N/A |
| cc_hearingloss | Self-reported severe hearing loss at baseline | N/A |
| cc_kidney_or_bladder_stones | Self-reported kidney or bladder stones at baseline | N/A |
| cc_arrhythmia | Self-reported arrhythmia at baseline | N/A |
| cc_gallbladder | Self-reported gallbladder disease at baseline | N/A |
| cc_tinnitus | Self-reported tinnitus at baseline | N/A |
| cc_ibd | Self-reported irritable bowel disorder at baseline | N/A |
| cc_osteopenia | Self-reported osteopenia at baseline | N/A |
| cc_nonmelanoma | Self-reported non-melanoma skin cancer at baseline | N/A |
| cc_osteoporosis | Self-reported osteoporosis at baseline | N/A |
| cc_hemorrhoids | Self-reported hemorrhoids at baseline | N/A |
| cc_gout | Self-reported gout at baseline | N/A |
| cc_glaucoma | Self-reported glaucoma at baseline | N/A |
| cc_bph | Self-reported benign prostatic hyperplasia at baseline | N/A |
| cc_diverticulosis | Self-reported diverticulosis at baseline | N/A |
| cc_pud | Self-reported peptic ulcers at baseline | N/A |
| cc_melanoma | Self-reported melanoma skin cancer at baseline | N/A |
| cc_diverticulitis | Self-reported diverticulitis at baseline | N/A |
| cc_hxmi | Self-reported myocardial infarction at baseline | N/A |
| cc_breast_cancer | Self-reported breast cancer at baseline | N/A |
| cc_copd_emphysema | Self-reported COPD (with emphysema) at baseline | N/A |
| cc_psoriasis | Self-reported psoriasis at baseline | N/A |
| cc_cad | Self-reported coronary artery disease (including angina) at baseline | N/A |
| cc_fibromyalgia | Self-reported fibromyalgia at baseline | N/A |
| cc_ra | Self-reported rheumatoid arthritis at baseline | N/A |
| cc_pe_or_dvt | Self-reported PE or DVT at baseline | N/A |
| cc_epilepsy | Self-reported epilepsy | N/A |
| cc_hashimotos | Self-reported Hashimoto’s disease | N/A |
| cc_pvd | Self-reported peripheral vascular disease | N/A |
| cc_prostate_cancer | Self-reported prostate cancer | N/A |
| cc_nonalcoholfattyliverdx | Self-reported non-alcoholic fatty liver disease | N/A |
| cc_thyroidgoiter | Self-reported goiter | N/A |
| cc_hepc | Self-reported Hepatitis C | N/A |
| cc_dm1 | Self-reported Diabetes type 1 | N/A |
| cc_macular_degeneration | Self-reported macular degeneration | N/A |
| cc_stroke | Self-reported stroke | N/A |
| cc_afib | Self-reported atrial fibrillation | N/A |
| cc_tia | Self-reported transient ischemic attack | N/A |
| cc_hepb | Self-reported Hepatitis B | N/A |
| **Physical Health-Related Symptoms** | | |
| stiffness | Self-reported stiffness at baseline | N/A |
| muscle_or_joint_pain | Self-reported muscle or joint pain at baseline | N/A |
| nasal_stuffiness | Self-reported nasal stuffiness at baseline | N/A |
| runny_nose | Self-reported runny nose at baseline | N/A |
| urination_at_night | Self-reported urination at night at baseline | N/A |
| floaters | Self-reported floaters at baseline | N/A |
| joint_pain_swelling | Self-reported joint pain swelling at baseline | N/A |
| itching_skin | Self-reported itching at baseline | N/A |
| cough | Self-reported cough at baseline | N/A |
| dryness | Self-reported dryness (skin) at baseline | N/A |
| easy_bruising_or_bleeding | Self-reported easy bruising or bleeding at baseline | N/A |
| tingling_or_numbness_in_extremities | Self-reported tingling or numbness in extremities at baseline | N/A |
| tingling_or_pins_and_needles | Self-reported tingling or pins and needles at baseline | N/A |
| heartburn | Self-reported heartburn at baseline | N/A |
| frequency_of_urination | Self-reported frequency of urination at baseline | N/A |
| constipation | Self-reported constipation at baseline | N/A |
| leg_cramps | Self-reported leg cramps at baseline | N/A |
| diarrhea | Self-reported diarrhea at baseline | N/A |
| ear_ringing | Self-reported ear ringing at baseline | N/A |
| heat_or_cold_intolerance | Self-reported heat or cold intolerance at baseline | N/A |
| night_sweats | Self-reported night sweats at baseline | N/A |
| dry_mouth | Self-reported dry mouth at baseline | N/A |
| excessive_belching_or_passing_of_gas | Self-reported excessive belching or Passing of gas at baseline | N/A |
| shortness_of_breath_with_exercise | Self-reported shortness of breath with exercise at baseline | N/A |
| memory_change | Self-reported memory change at baseline | N/A |
| lightheadedness | Self-reported lightheadedness at baseline | N/A |
| sinus_pain | Self-reported sinus pain at baseline | N/A |
| shortness_of_breath | Self-reported shortness of breath at baseline | N/A |
| swelling_in_calves_or_feet | Self-reported swelling in calves or feet at baseline | N/A |
| coughing_up_sputum | Self-reported coughing up sputum at baseline | N/A |
| urgency | Self-reported urgency at baseline | N/A |
| hay_fever | Self-reported hay fever at baseline | N/A |
| discharge | Self-reported discharge (nose and sinuses) at baseline | N/A |
| hemorrhoids | Self-reported hemorrhoids at baseline | N/A |
| cramping | Self-reported cramping at baseline | N/A |
| numbness_or_loss_of_sensation | Self-reported numbness or loss of sensation at baseline | N/A |
| fa_ind | Self-reported food allergies (any vs. none, ignoring additional details about which allergen) | N/A |
| sa_ind | Self-reported seasonal allergies (any vs. none, ignoring additional details about which allergen) | N/A |
| nsa_ind | Self-reported non-seasonal allergies (any vs. none, ignoring additional details about which allergen) | N/A |
| ma_ind | Self-reported medication allergies (any vs. none, ignoring additional details about which allergen) | N/A |

**A.4. On Site Assessments - Mental Health Surveys**

| **Variable Name** | **Definition** | **Description of Derivation** |
| --- | --- | --- |
| gad7_total_score | Generalized Anxiety Disorder-7 total score (range 0, 21) at baseline | Sum of 7 individual questions  For analysis:  = log(gad7_total_score + 1) |

**A.5. Blood Draw - Standard Laboratory Data**

| **Variable Name** | **Definition** | **Description of Derivation** |
| --- | --- | --- |
| standard_labs_hemoglobin_gdl | Hemoglobin (g/dl) at baseline | N/A |
| standard_labs_serum_creatinine_mgdl | Serum Creatinine (mg/dl) at baseline | N/A |
| standard_labs_hdl_mgdl | High density lipoprotein (mg/dl) at baseline | N/A |
| standard_labs_ldl_mgdl | Low density lipoprotein (mg/dl) at baseline | N/A |
| standard_labs_triglycerides_mgdl | Triglycerides (mg/dl) at baseline | N/A |
| standard_labs_hba1c_pct_tl_hgb | Hemoglobin A1c (%) at baseline | N/A |
| standard_labs_alt_ul | Alanine aminotransferase (U/L) at baseline | N/A |
| standard_labs_ast_ul | Aspartate aminotransferase (U/L) at baseline | N/A |
| standard_labs_vitamind_ngml | Vitamin D (ng/ml) at baseline | N/A |
| standard_labs_crp_mgl | C-reactive protein (mg/l) at baseline | N/A |
| standard_labs_blood_glucose_mgdl | Blood glucose (mg/dl) at baseline | N/A |
| standard_labs_neutrophil_segs_pct_wbc | Neutrophil segments (% WBC) at baseline | N/A |
| standard_labs_neutrophils_thou_per_mcl | Total neutrophils (k/mcL) at baseline | N/A |
| standard_labs_lymphocytes_thou_per_mcl | Total lymphocytes (k/mcL) at baseline | N/A |
| standard_labs_magnesium_meql | Magnesium (MEQ/L) at baseline | N/A |
| standard_labs_monocytes_thou_per_mcl | Absolute Monocytes (k/mcL) at baseline | N/A |
| standard_labs_eosinophils_thou_per_mcl | Absolute Eosinophils (k/mcL) at baseline | N/A |
| standard_labs_basophils_thou_per_mcl | Absolute Basophils (k/mcL) at baseline | N/A |
| standard_labs_hematocrit_pct_rbc_blood | Hematocrit (% RBC to whole blood volume) at baseline | N/A |
| standard_labs_mcv_fl | Mean corpuscular volume (fL) at baseline | N/A |
| standard_labs_mch_pg | Mean corpuscular hemoglobin (pg) at baseline | N/A |
| standard_labs_mpv_fl | Mean platelet volume (fL) at baseline | N/A |
| standard_labs_platelets_per_cumm | Platelet count (cumm) at baseline | N/A |
| standard_labs_rbc_mill_per_mcl | Red blood cell count (millions/mcL) at baseline | N/A |
| standard_labs_wbc_thou_per_mcl | White blood cell count (millions/mcL) at baseline | N/A |
| standard_labs_calcium_mgdl | Calcium (mg/dL) at baseline | N/A |
| standard_labs_cholesterol_mgdl | Total cholesterol (mg/dL) at baseline | N/A |
| standard_labs_chloride_meql | Chloride (MEQ/L) at baseline | N/A |
| standard_labs_potassium_meql | Potassium (MEQ/L) at baseline | N/A |
| standard_labs_sodium_meql | Sodium (MEQ/L) at baseline | N/A |
| standard_labs_protein_serum_gdl | Protein in serum (g/dL) at baseline | N/A |
| standard_labs_albumin_gdl | Albumin (g/L) at baseline | N/A |
| standard_labs_uric_acid_mgdl | Uric acid (mg/dL) at baseline | N/A |
| standard_labs_creatinine_random_urine_mgl | Creatinine in urine (mg/dL) at baseline | N/A |
| standard_labs_gfr_mdrd_ml_min | Glomerular filtration rate (mL/min/1.73 m^2^) based on Modification of Diet in Renal Disease Study equation at baseline | N/A |
| standard_labs_reticulocytes_bill_per_liter | Absolute reticulocytes (billions/L) at baseline | N/A |
| standard_labs_tsh_miu_per_liter | Thyroid stimulating hormone (mIU/L) at baseline | N/A |
| standard_labs_specific_gravity | Urine specific gravity at baseline | N/A |
| standard_labs_reaction_ph | Urine reaction pH at baseline | N/A |

**A.6. Participant Portal (App) Surveys**

**SES-Related Variables (Life Circumstances and Habits Survey)**

| **Variable Name** | **Definition** | **Description of Derivation** |
| --- | --- | --- |
| educ_hs_or_less | Self-reported highest education level completed from first survey completed | N/A |
| inc_under_25 | Self-reported household income from first survey completed | N/A |
| marital_married | Self-reported marital status from first survey completed | N/A |
| employ_not_working | Self-reported employment status from first survey completed | N/A |
| insured_no | Self-reported health insurance (yes/no) from first survey completed | N/A |

**A.7. Sensors Data**

| **Variable Name** | **Definition** | **Description of Derivation** |
| --- | --- | --- |
| mean_steps_first_30_days | Average daily number of steps in the first 30 days in study (measured with study watch) | Daily step count in first 30 days / 30 days |
| mean_days_10hrs_wear_first_30days | Number of days of 10 hours of wear in the first 30 days in study (measured with study watch) |  |
